# Prediction of Pulmonary Vein Isolation and Gap Recurrence on 12-Lead ECG Using Deep Learning

**DOI:** 10.1101/2025.11.24.25340932

**Authors:** Iris Liu, Meichen Liu, Isabelle Ah-Sen, Wenteng Hou, Habib R. Khan, Christopher Cheung, Lorne Gula, Anthony Tang, Jaimie Manlucu, Peter Leong-Sit, Allan Skanes, Raymond Yee, Pavel Antiperovitch

## Abstract

**Background:** Pulmonary vein isolation (PVI) is key to atrial fibrillation (AF) ablation, but arrhythmia often recurs due to conduction gaps permitting pulmonary vein (PV) reconnection. Currently, gap identification requires invasive remapping. We evaluated whether deep learning applied to surface electrocardiograms (ECGs) could (i) detect the electrophysiologic signature of PVI and (ii) predict PV reconnection at redo ablation.

**Methods:** We retrospectively studied 176 patients (2012–2023) who had initial PVI and repeat ablation. A total of 865 10-second 12-lead ECGs were extracted from GE MUSE and CardioLab systems, segmented into 1-2 second clips, and used to train ResNet-based convolutional neural networks. Separate models were developed for: (i) PVI detection (pre-vs. post-ablation ECGs) and (ii) gap prediction using pre-redo ECGs. Demographic features were tested alone and in multimodal fusion with ECGs. Performance was evaluated using Receiver Operating Characteristic (ROC) curves, sensitivity, and specificity with stratified cross-validation. Gradient-weighted class activation mapping (Grad-CAM) assessed feature importance.

**Results:** The best PVI detection model distinguished ECGs before and after PVI with the area under the ROC (AUROC) = 0.879. Grad-CAM localized attention to the diastolic period and P-wave morphology. For gap prediction, the model trained on pre-redo ECGs achieved an AUROC of 0.819 (sensitivity 77.5%, specificity 75.8%). Adding demographics improved the AUROC to 0.830 (sensitivity 84%, specificity 72%), whereas demographics alone performed no better than chance (AUROC = <50%). Feature importance analysis highlighted P-wave onset and offset, inter-ablation time interval, left atrial volume index, age, and left ventricular ejection function as the strongest contributors in gap prediction.

**Conclusions:** Deep learning identifies a consistent ECG biosignature of acute PVI and predicts PV reconnection before redo ablation with moderate accuracy, primarily using P-wave morphology. These models may inform patient selection, procedural planning, and counselling in patients with recurrent AF after PVI.

**GRAPHICAL ABSTRACT:** 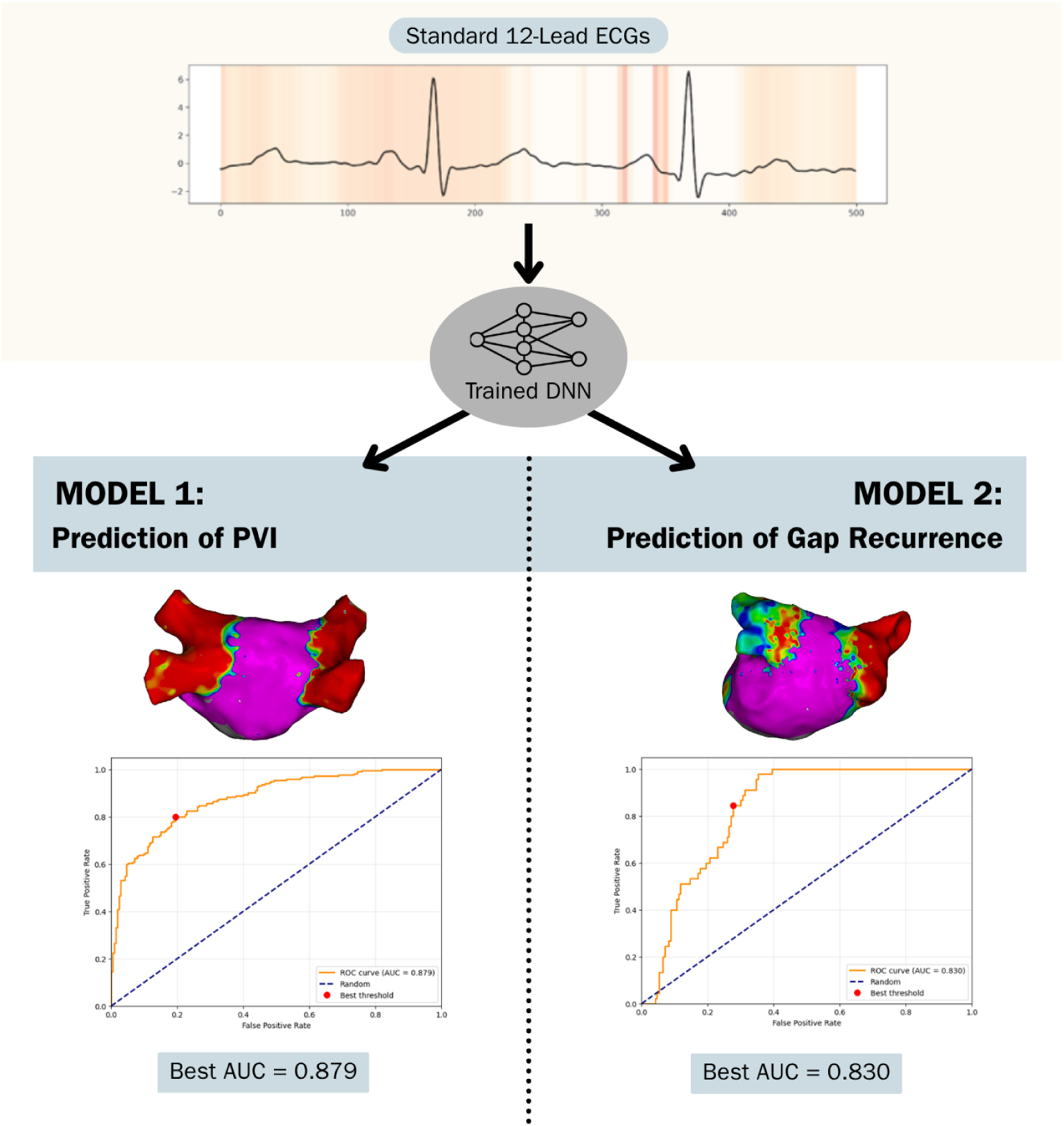

**CLINICAL PERSPECTIVE:** *What is Known:*

- Arrhythmia recurrence post-pulmonary vein isolation (PVI) are commonly caused by conduction gaps in the PV, and repeat isolation procedures lead to better arrhythmia-free survival compared to if recurrences are due to extrapulmonary triggers with chronically isolated veins.
- Currently, there are no non-invasive methods to detect conduction gaps after PVI.

*What the Study Adds:*

- Our deep learning models can predict the presence of conduction gaps from non-invasive surface electrocardiograms prior to repeat procedures with moderate accuracy.
- We identified important ECG and demographic features in predicting gap presence.
- Ultimately, we provide potential methods of risk-stratifying patients and selecting candidates for repeat procedures that can be accessed in outpatient settings.

## INTRODUCTION

Atrial fibrillation (AF) is the most common sustained cardiac arrhythmia and is associated with impaired quality of life as well as increased morbidity and mortality.^1^ Pulmonary vein isolation (PVI) is the cornerstone catheter ablation strategy that involves electrically isolating the pulmonary veins (PV) to treat AF triggers. Randomized trials such as EARLY-AF have demonstrated the superiority of ablation over pharmacotherapy in maintaining sinus rhythm.^2^ Despite procedural success, arrhythmia recurs in ∼30% of patients, often due to pulmonary vein (PV) reconnection through gaps or other non-PV triggers.^3,4^ Pulmonary vein reconnection identified at repeat ablation, when successfully re-isolated, is associated with favourable long-term outcomes.^5,6^ In contrast, AF recurrence despite durable PV isolation suggests non-PV substrate that may be less amenable to further ablation^7–10^.

Currently, gap identification requires invasive remapping at redo procedures. Empiric ablation in the absence of gaps has not demonstrated a consistent benefit outside of specialized mapping techniques like TAILORED-AF.^7^ A noninvasive method to detect PV reconnection before or after ablation could guide patient selection, procedural planning, and follow-up strategies.

Deep learning, particularly convolutional neural networks (CNNs), excels at detecting complex spatial and temporal patterns in large datasets.^11^ Prior work with AF has attempted to predict recurrence after PVI using structural imaging (e.g. 3D left atrial reconstruction) with modest accuracy (AUC ∼0.61).^12^ Traditional ECG analyses have suggested that p-wave indices, such as dispersion or filtered p-wave duration, change after ablation and may correlate with recurrence, but these markers lack consistent independent predictive value.^13–15^ Thus, while P-wave morphology may encode signatures of atrial remodelling and PV isolation, deep learning applied directly to raw 12-lead ECGs has not been systematically studied in this context.

We therefore sought to develop and validate AI models capable of: (i) detecting the electrophysiological biosignature of PVI on standard surface ECGs, (ii) predicting conduction gaps in previously ablated PVI lines prior to redo procedures, and (iii) evaluating whether baseline ECG and clinical features could stratify the risk of PV reconnection before initial ablation.

## METHODS

### Patient Selection

This is a retrospective observational study that collected data on patients with atrial fibrillation who underwent an initial ablation procedure for AF and one repeat ablation procedure for recurrent AF or atrial flutter. We screened electrophysiology procedure records dated between January 1, 2012 to December 31, 2023 at University Hospital in London, Ontario, Canada. Patients with missing procedural clinical records or intra-procedural rhythm tracings were excluded. The study was approved by Western University Research Ethics Board and Lawson Research Institute (HSREB reference: 2024-124520-90091).

### Data Collection

Patient demographics, comorbidities, medication regimens, and procedure details, including ablation lines performed and subsequent isolation results, were extracted from patient electronic charts (see Table 1 for a summary of collected information). Details regarding presence of conduction gaps on repeat ablation were manually gleaned from procedure reports. We collected raw ECG signal data from two types of 12-lead ECGs: Outpatient ECGs (ECG OP) from the GE MUSE platform (Chicago, Illinois), recorded either in clinic or in an ambulatory setting in a hospital, and intraprocedural 12-lead ECGs (ECG IP) from GE CardioLab. Each ECG record contained a 10-second segment of the raw ECG signal sampled at 500 Hz.

**Table 1.**
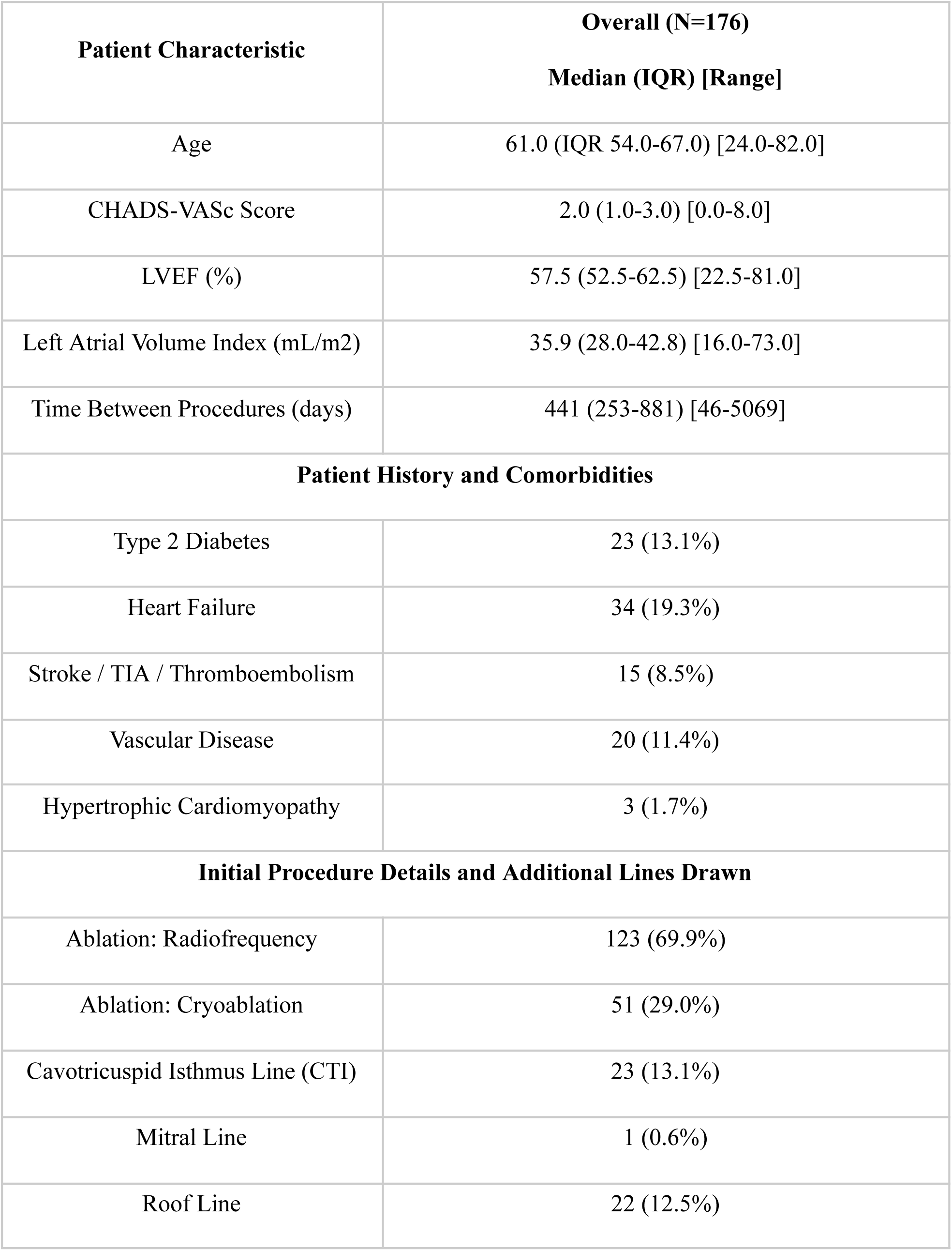
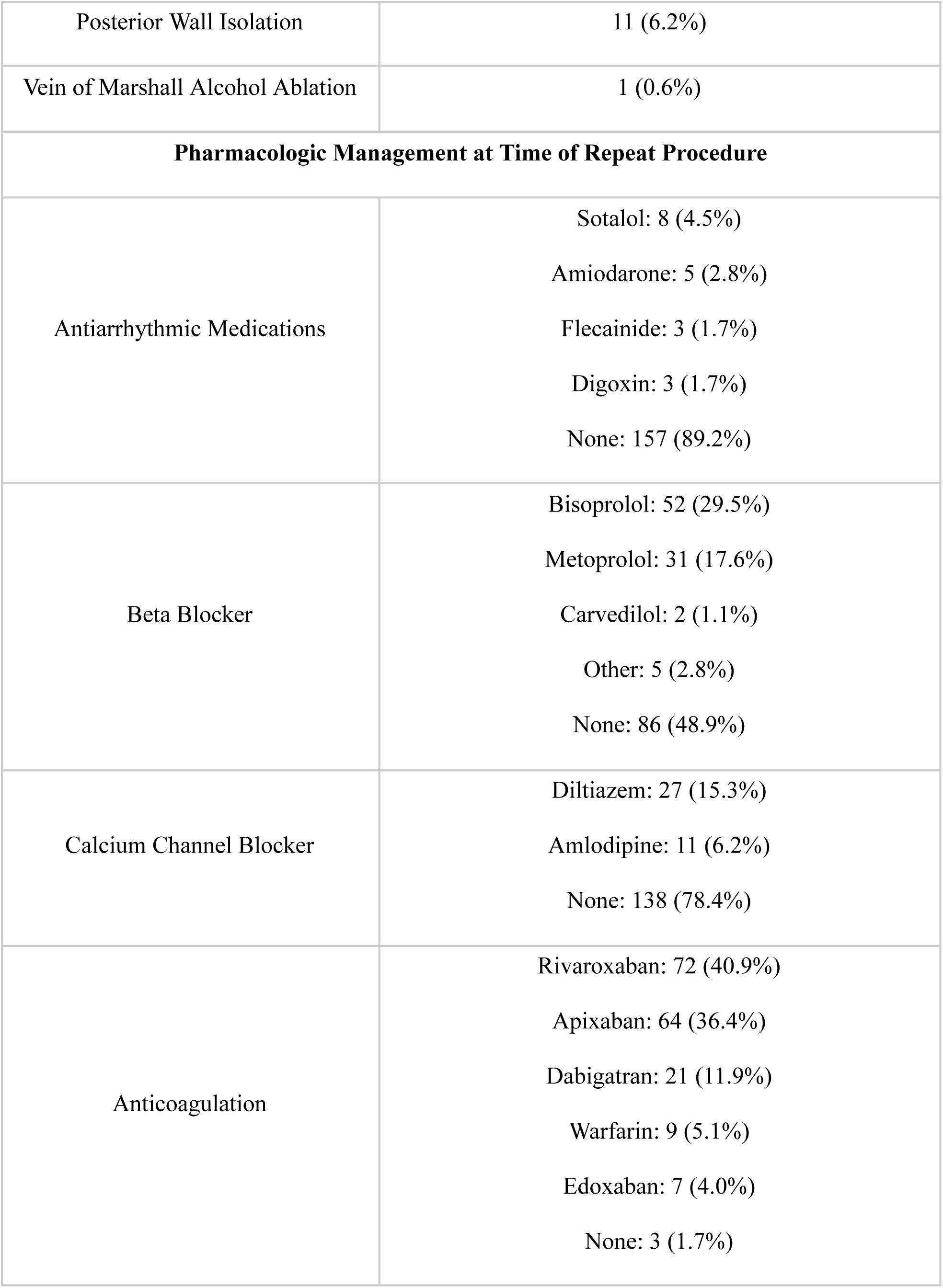
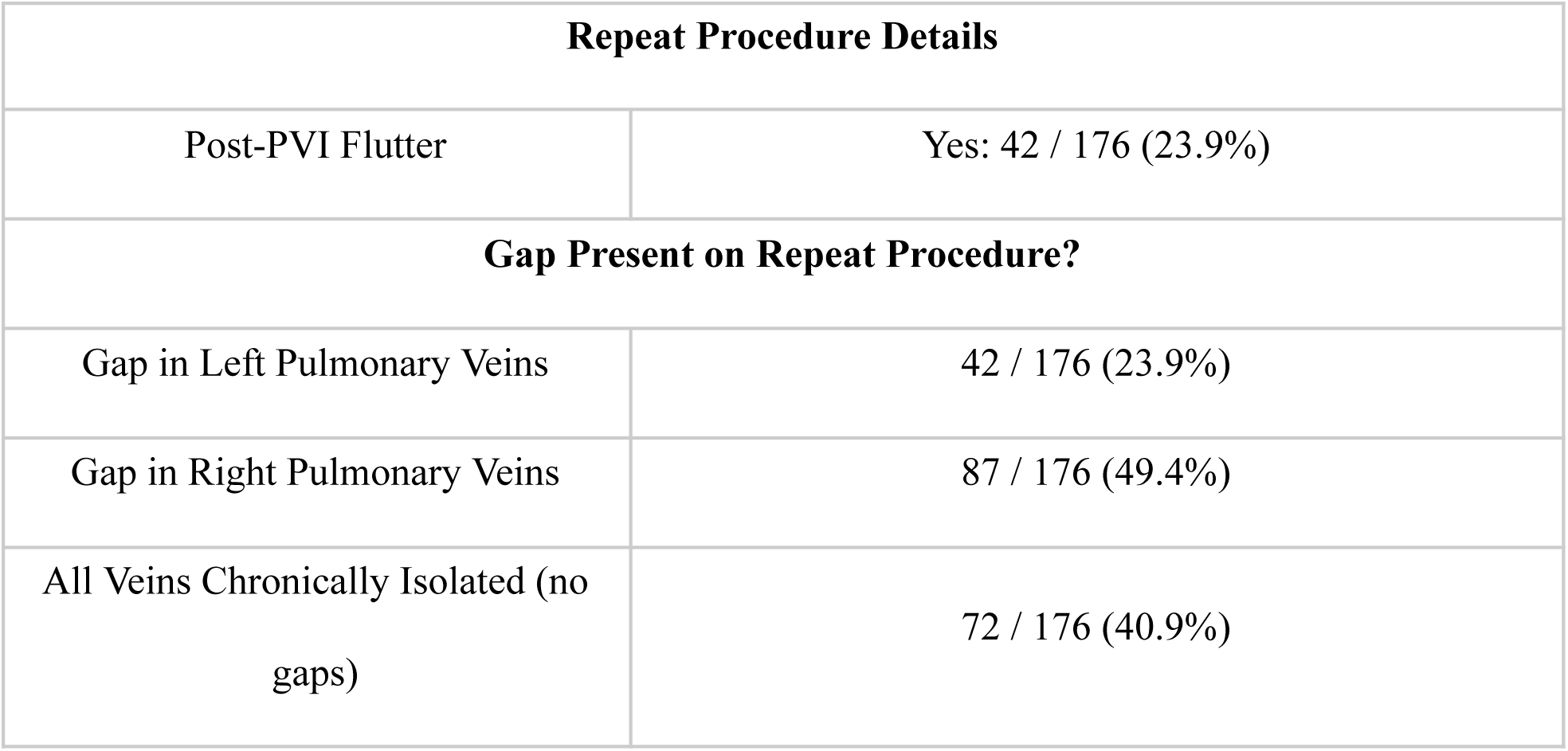
Summary of baseline participant demographics and redo ablation details. Numerical variables are expressed as median, interquartile range (IQR), and total range. Categorical variables are summarized as sub-category counts.

| **Patient Characteristic** | **Overall (N=176)**<br>**Median (IQR) [Range]** |
| --- | --- |
| Age | 61.0 (IQR 54.0-67.0) [24.0-82.0] |
| CHADS-VASc Score | 2.0 (1.0-3.0) [0.0-8.0] |
| LVEF (%) | 57.5 (52.5-62.5) [22.5-81.0] |
| Left Atrial Volume Index (mL/m2) | 35.9 (28.0-42.8) [16.0-73.0] |
| Time Between Procedures (days) | 441 (253-881) [46-5069] |
| **Patient History and Comorbidities** |  |
| Type 2 Diabetes | 23 (13.1%) |
| Heart Failure | 34 (19.3%) |
| Stroke / TIA / Thromboembolism | 15 (8.5%) |
| Vascular Disease | 20 (11.4%) |
| Hypertrophic Cardiomyopathy | 3 (1.7%) |
| **Initial Procedure Details and Additional Lines Drawn** |  |
| Ablation: Radiofrequency | 123 (69.9%) |
| Ablation: Cryoablation | 51 (29.0%) |
| Cavotricuspid Isthmus Line (CTI) | 23 (13.1%) |
| Mitral Line | 1 (0.6%) |
| Roof Line | 22 (12.5%) |
| Posterior Wall Isolation | 11 (6.2%) |
| Vein of Marshall Alcohol Ablation | 1 (0.6%) |
| **Pharmacologic Management at Time of Repeat Procedure** |  |
| Antiarrhythmic Medications | Sotalol: 8 (4.5%)<br>Amiodarone: 5 (2.8%)<br>Flecainide: 3 (1.7%)<br>Digoxin: 3 (1.7%)<br>None: 157 (89.2%) |
| Beta Blocker | Bisoprolol: 52 (29.5%)<br>Metoprolol: 31 (17.6%)<br>Carvedilol: 2 (1.1%)<br>Other: 5 (2.8%)<br>None: 86 (48.9%) |
| Calcium Channel Blocker | Diltiazem: 27 (15.3%)<br>Amlodipine: 11 (6.2%)<br>None: 138 (78.4%) |
| Anticoagulation | Rivaroxaban: 72 (40.9%)<br>Apixaban: 64 (36.4%)<br>Dabigatran: 21 (11.9%)<br>Warfarin: 9 (5.1%)<br>Edoxaban: 7 (4.0%)<br>None: 3 (1.7%) |

| Repeat Procedure Details |  |
| --- | --- |
| Post-PVI Flutter | Yes: 42 / 176 (23.9%) |
| Gap Present on Repeat Procedure? |  |
| Gap in Left Pulmonary Veins | 42 / 176 (23.9%) |
| Gap in Right Pulmonary Veins | 87 / 176 (49.4%) |
| All Veins Chronically Isolated (no gaps) | 72 / 176 (40.9%) |

### Machine Learning Analysis of Demographic Data

Demographic data including baseline characteristics and comorbidities, were analyzed using the packages Numpy, Scikit-learn, XGBoost, and PyTorch in Python 3.10.12. We applied a wide range of machine learning algorithms to optimize predictive ability, including regularized logistic regression, Random Forest, and XGBoost.

Randomized stratified k-fold cross-validation was used to select the best hyperparameters in these models. Numerical features were imputed as median values whereas categorical features were hot encoded, and missing variables were recorded as additional categories for categorical variables. All patients had valid target variables. Allowable predictors for each outcome were predetermined. In each experiment, the models were fit to a training dataset (75%) and evaluated on an independent testing set (25%). Model performance was assessed using area under the receiver operating characteristic curve (AUROC), sensitivity, and specificity computed on the testing set. Average area under the curve (AUC) and 95% confidence intervals (CI) were calculated across the 500 runs using the percentile method. Sensitivity and specificity were determined according to the best threshold on the AUC curve. Feature importance, which indicates better predictors of long-term PV isolation, was also identified using Shapley evaluations.

### Deep Learning on ECG Signal Data

A ResNet convolutional neural network (CNN) architecture was employed for deep learning on ECG signal data.^16^ The models were hosted on a GPU-enabled cluster at Western University, utilizing Python 3.10.12 and PyTorch 2.4.0 with an H100 GPU.

### Signal Pre-processing

To augment the number of training samples, the raw ECG signals were segmented into 2-second clips. All 12-lead surface ECGs were sampled at 250 Hz, and intracardiac signals were resampled at 500 Hz. We developed an automated preprocessing pipeline to standardize the datasets. First, Outlier signals were cropped at a specific range after centering around the mean. Then, artifacts, such as those caused by A/C-induced 60 Hz interference, were detected when their amplitude exceeded two standard deviations (SDs) above the local mean. These artifacts were flattened using a local moving average curve, with a window size of 10. Finally, amplitude was normalized leadwise to have a mean of 0 and a standard deviation of 1.

### Model Architecture and Training

ResNet served as the foundational CNN backbone for modelling the ECGs. Features extracted by ResNet were subsequently combined with a baseline multilayer perceptron (MLP) to integrate the predictive capabilities of both ECG and demographic features. The dataset was randomly split into training and testing sets with a 75/25 ratio. Hyperparameter tuning was performed using randomized stratified k-fold cross-validation for all models. Tuned parameters include filter size, number of filters, dropout rate, number of blocks built in the CNN, learning rate, batch size, number of epochs, and hidden dimensions of the MLP for demographic features. Gradient-weighted Class Activation Mapping (Grad-CAM) heat maps were used for model explainability.^17^

## RESULTS

We screened 250 patients who underwent the two procedures during the study time period, and 176 patients met the inclusion criteria. Seventy-four patients were excluded due to missing procedure notes or signal data records. All patients underwent two ablation procedures, which allowed us to evaluate the presence of gaps after the initial procedure. The baseline demographics of the included patients are outlined in Table 1. The cohort represents the standard PVI population with median age of 61.0 (IQR 54.0-67.0) and a wide range of left ventricular ejection fractions (LVEF 22.5-81.0%) and left atrial volume indexes (LAVI 16.0-73.0 mL/m^2^). All PVI was performed using radiofrequency or cryotherapy ablation.

### Identifying PVI on ECG

Our first objective was to determine whether a deep neural network (DNN) could identify electrophysiologic evidence of pulmonary vein isolation (PVI) on surface ECGs. A model trained on 10-second 12-lead ECGs obtained immediately before and after the procedure (DNN-PVI1-10s) successfully distinguished pre- and post-PVI recordings, achieving an AUROC of 0.784 (SD 0.069). To reduce input complexity and increase the number of training examples, each tracing was augmented by segmentation into single-cycle one-second clips centered on the QRS complex (DNN-PVI1-1s). This approach substantially improved performance, with an AUROC of 0.866 (SD 0.017) (Table 2). To test generalizability, we trained an additional model on outpatient ECGs obtained outside of the procedural setting (DNN-PVI1-OP-1s). While performance was slightly attenuated, the model still demonstrated predictive ability, with an AUROC of 0.771 (SD 0.016). The best performance overall was observed in distinguishing outpatient ECGs acquired before the first ablation from those after the second ablation procedure, yielding an AUROC of 0.879 (SD 0.016) (Figure 1A). Model interpretability analysis using Grad-CAM demonstrated that network attention was primarily directed to the diastolic interval, with secondary weighting toward P-waves (Figure 2).

**Figure 1.**
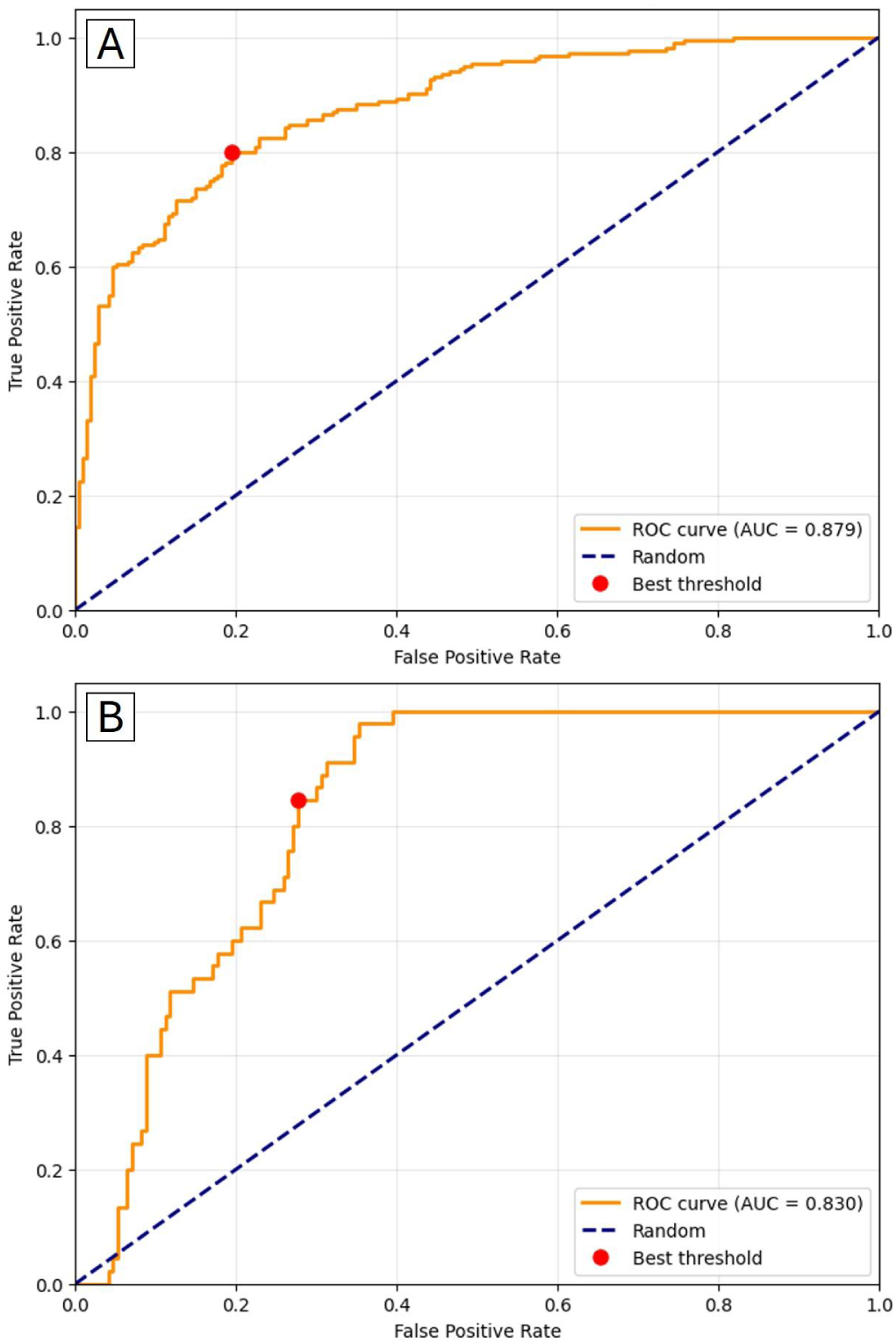
Performance of Deep Learning Models for PVI Detection and Gap Prediction. (A) Receiver operating characteristic (ROC) curve for distinguishing pre-versus post-pulmonary vein isolation (PVI) using the best-performing model trained on outpatient ECGs before the first and after the second ablation (DNN-PVI2-OP-2s; AUROC 0.879). (B) ROC curve for predicting the presence of conduction gaps at repeat ablation using 2s pre-procedure outpatient ECG segments combined with baseline demographic features (DNN-Gap-OP-Demo-2s; AUROC 0.830).

**Figure 2.**
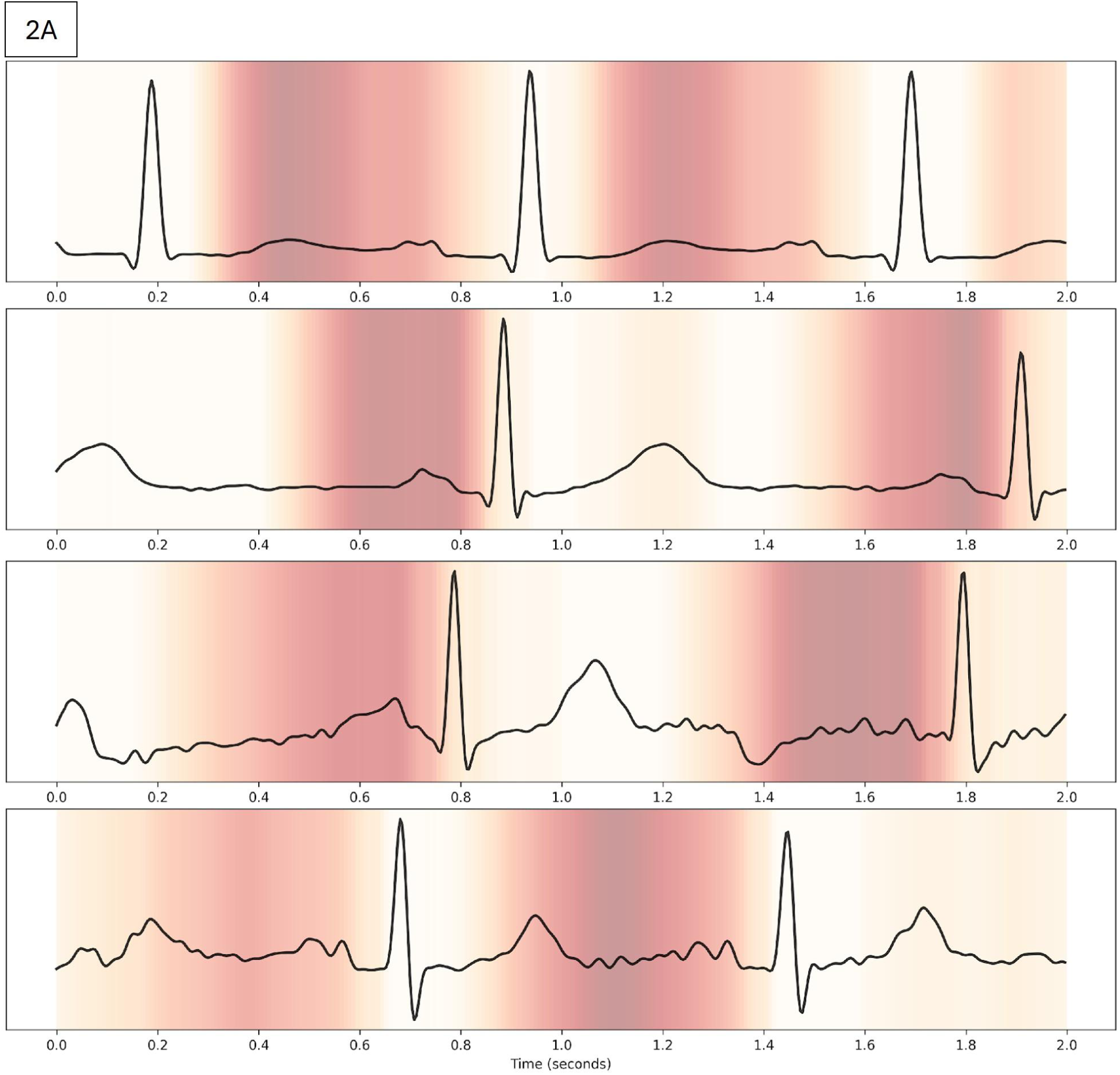
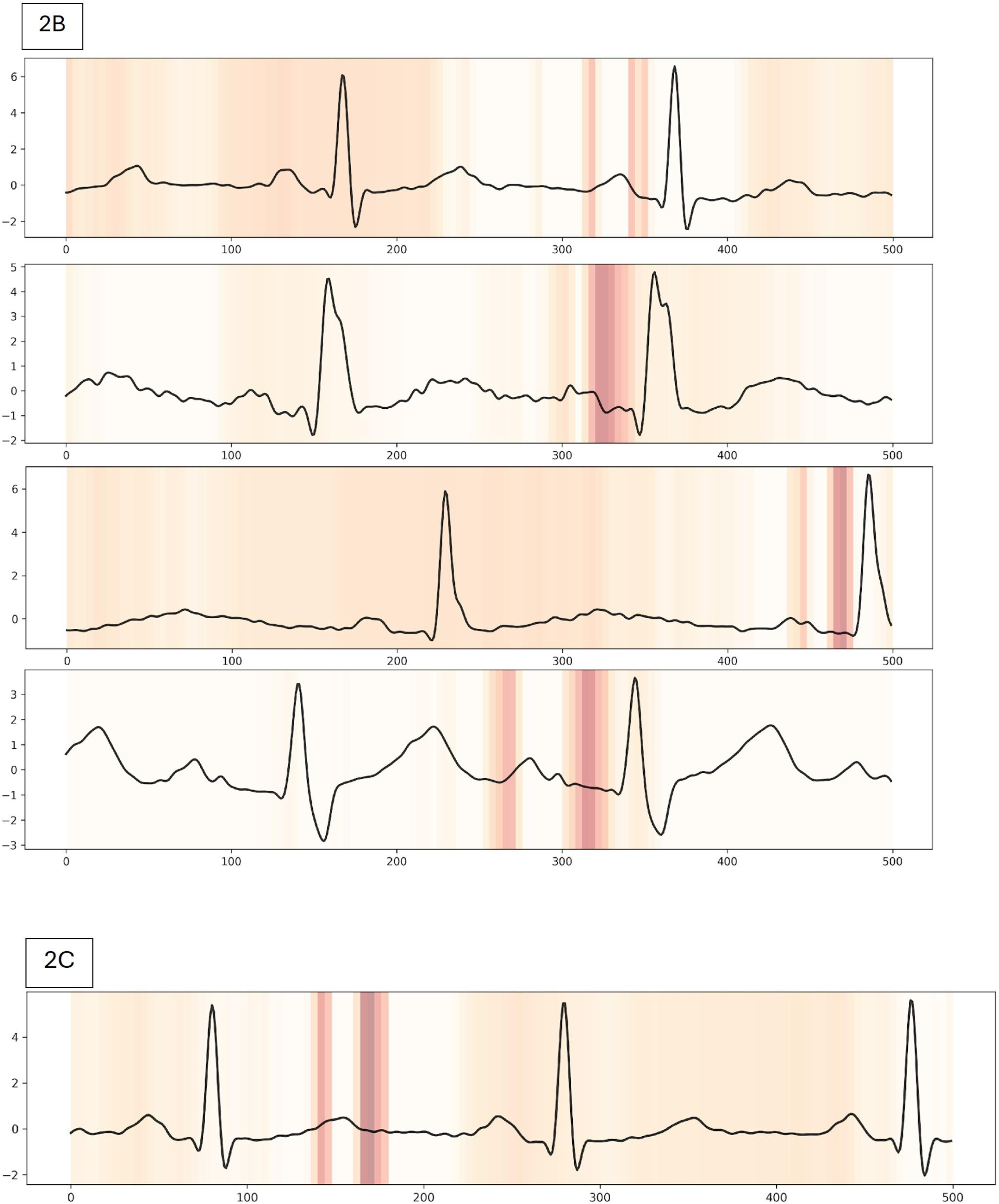
Grad-CAM Interpretability of Deep Learning Models. (A) Representative Grad-CAM overlays from four outpatient ECG tracings analyzed by the PVI detection model (DNN-PVI1-OP-2s). Network attention was predominantly focused on the diastolic interval, with secondary emphasis on P-wave morphology. (B) Representative Grad-CAM overlays from four outpatient ECG tracings analyzed by the gap-prediction model (DNN-Gap-OP-Demo-2s). Predictive features were localized to the onset and offset of the P-wave. (C) Example of model misclassification: a patient with a true conduction gap incorrectly predicted as “no gap”. Grad-CAM demonstrated misplaced attention on the T-wave rather than the P-wave region.

**Table 2.**
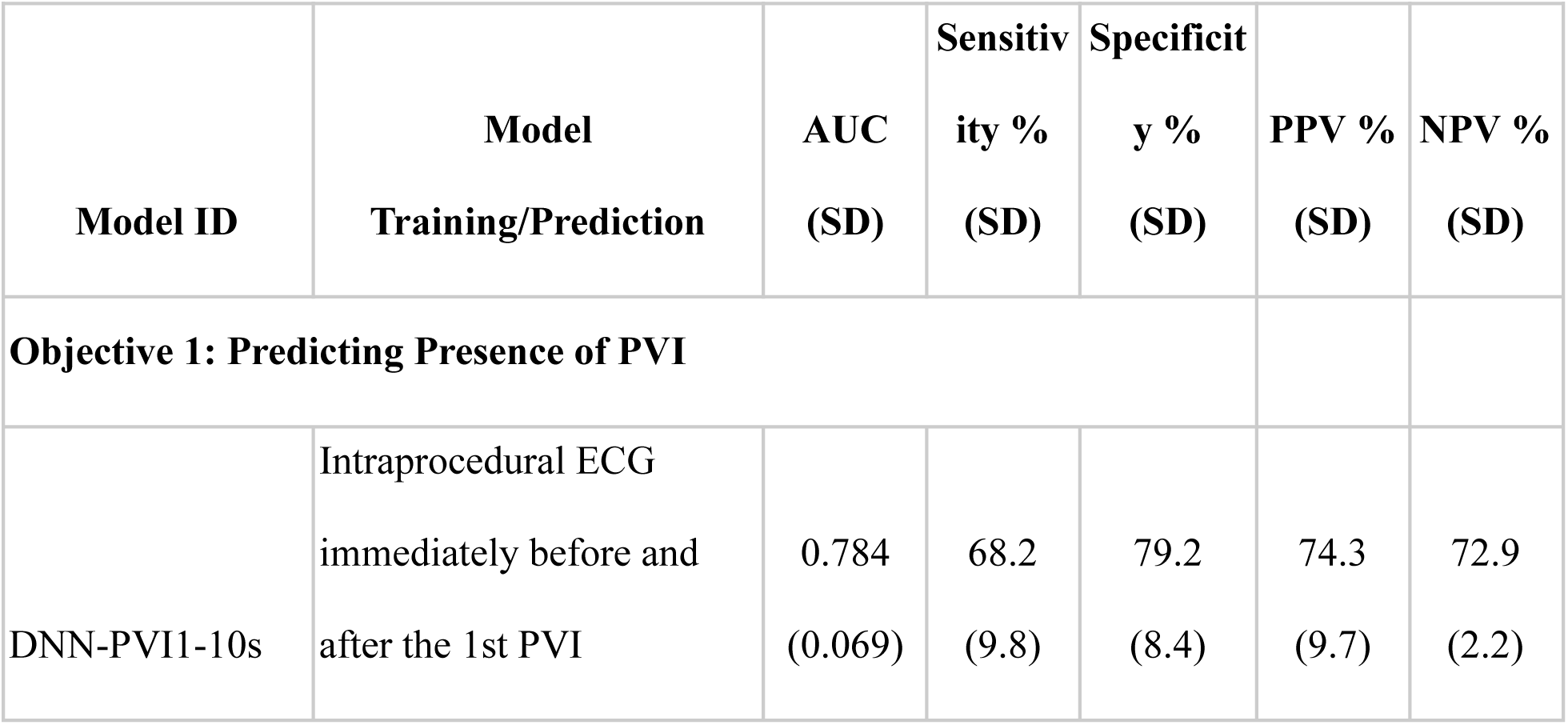
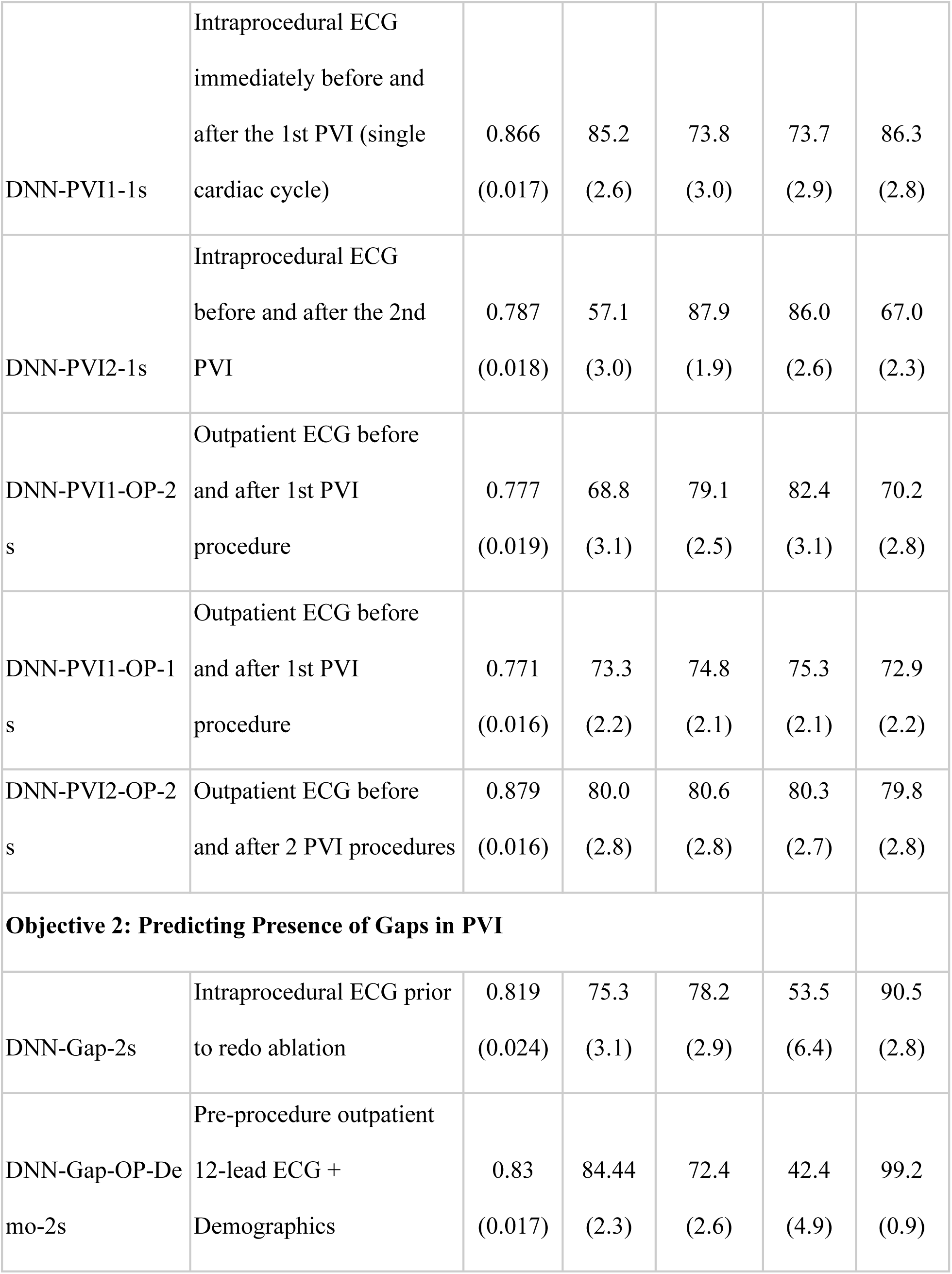
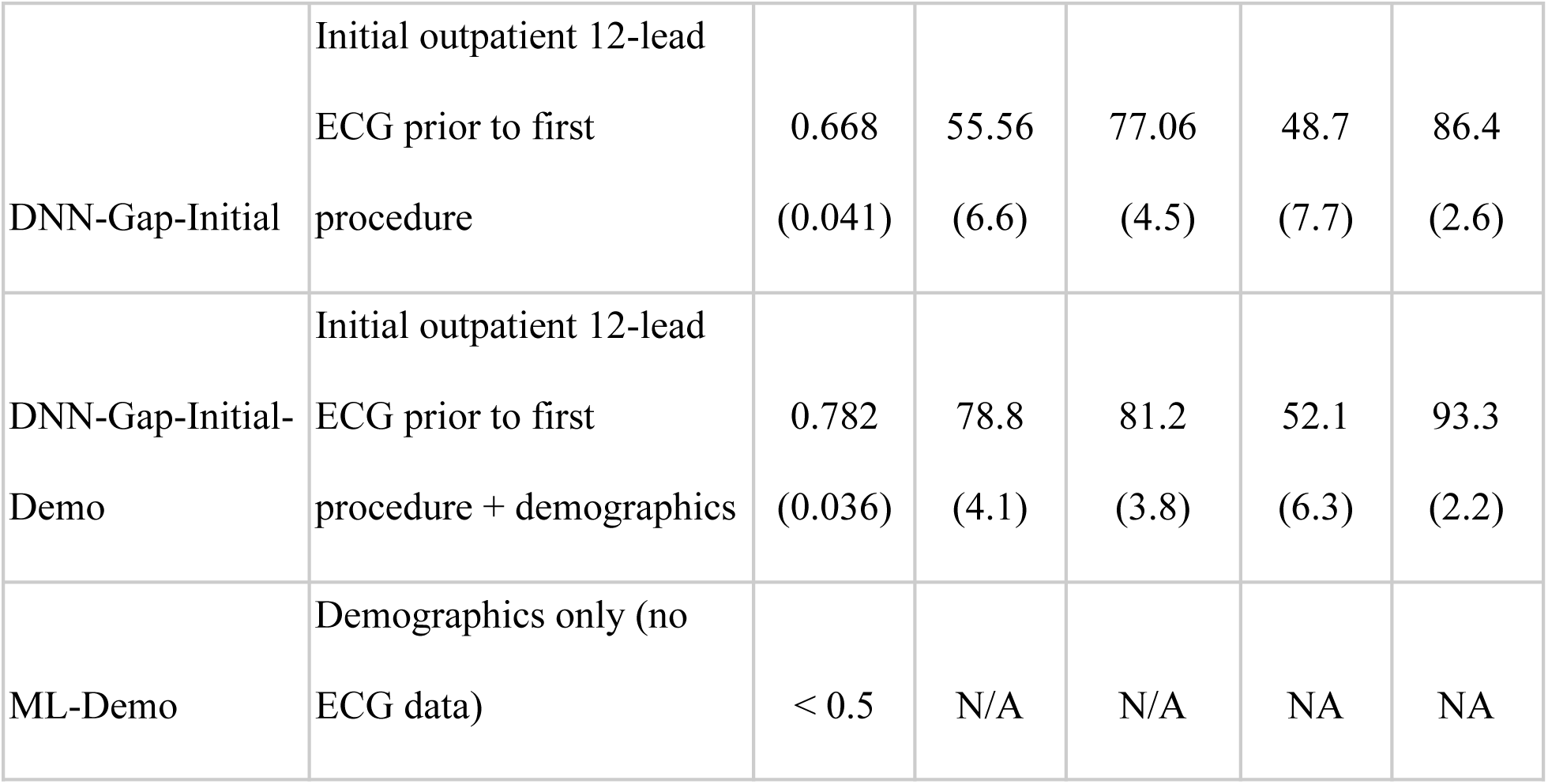
Performance of deep learning models for detecting pulmonary vein isolation and predicting conduction gaps. Data entities used to train each model version are listed, along with their respective AUROCs, sensitivities, specificity, positive predictive values (PPV), and negative predictive values (NPV).

| Model ID | Model<br>Training/Prediction | AUC<br>(SD) | Sensitivity %<br>(SD) | Specificity %<br>(SD) | PPV %<br>(SD) | NPV %<br>(SD) |
| --- | --- | --- | --- | --- | --- | --- |
| **Objective 1: Predicting Presence of PVI** |  |  |  |  |  |  |
| DNN-PVI1-10s | Intraprocedural ECG<br>immediately before and<br>after the 1st PVI | 0.784<br>(0.069) | 68.2<br>(9.8) | 79.2<br>(8.4) | 74.3<br>(9.7) | 72.9<br>(2.2) |
| DNN-PVI1-1s | Intraprocedural ECG immediately before and after the 1st PVI (single cardiac cycle) | 0.866<br>(0.017) | 85.2<br>(2.6) | 73.8<br>(3.0) | 73.7<br>(2.9) | 86.3<br>(2.8) |
| DNN-PVI2-1s | Intraprocedural ECG before and after the 2nd PVI | 0.787<br>(0.018) | 57.1<br>(3.0) | 87.9<br>(1.9) | 86.0<br>(2.6) | 67.0<br>(2.3) |
| DNN-PVI1-OP-2s | Outpatient ECG before and after 1st PVI procedure | 0.777<br>(0.019) | 68.8<br>(3.1) | 79.1<br>(2.5) | 82.4<br>(3.1) | 70.2<br>(2.8) |
| DNN-PVI1-OP-1s | Outpatient ECG before and after 1st PVI procedure | 0.771<br>(0.016) | 73.3<br>(2.2) | 74.8<br>(2.1) | 75.3<br>(2.1) | 72.9<br>(2.2) |
| DNN-PVI2-OP-2s | Outpatient ECG before and after 2 PVI procedures | 0.879<br>(0.016) | 80.0<br>(2.8) | 80.6<br>(2.8) | 80.3<br>(2.7) | 79.8<br>(2.8) |
| **Objective 2: Predicting Presence of Gaps in PVI** |  |  |  |  |  |  |
| DNN-Gap-2s | Intraprocedural ECG prior to redo ablation | 0.819<br>(0.024) | 75.3<br>(3.1) | 78.2<br>(2.9) | 53.5<br>(6.4) | 90.5<br>(2.8) |
| DNN-Gap-OP-Demo-2s | Pre-procedure outpatient 12-lead ECG + Demographics | 0.83<br>(0.017) | 84.44<br>(2.3) | 72.4<br>(2.6) | 42.4<br>(4.9) | 99.2<br>(0.9) |
| DNN-Gap-Initial | Initial outpatient 12-lead ECG prior to first procedure | 0.668<br>(0.041) | 55.56<br>(6.6) | 77.06<br>(4.5) | 48.7<br>(7.7) | 86.4<br>(2.6) |
| DNN-Gap-Initial-Demo | Initial outpatient 12-lead ECG prior to first procedure + demographics | 0.782<br>(0.036) | 78.8<br>(4.1) | 81.2<br>(3.8) | 52.1<br>(6.3) | 93.3<br>(2.2) |
| ML-Demo | Demographics only (no ECG data) | < 0.5 | N/A | N/A | NA | NA |

### Identifying Conduction Gaps in PVI

We next trained a model to predict the presence of conduction gaps in PVI using pre-procedural surface ECGs obtained at the start of the redo ablation procedure, with intracardiac mapping serving as the reference standard. A model trained on two-second ECG segments centered on the QRS complex (DNN-Gap-2s) achieved an AUROC of 0.819 (SD 0.024), corresponding to a sensitivity of 75.3% (±3.1) and specificity of 78.2% (±2.9). Incorporating demographic features (age, sex, comorbidities; see Table 1) into the model (DNN-Gap-OP-Demo-2s) yielded a modest improvement in discrimination, with an AUROC of 0.830 (SD 0.017, sensitivity=84%, specificity=72%), indicating incremental predictive value of baseline clinical characteristics (Figure 1B).

Feature importance analysis showed that the interval between ablation procedures, patient age, LAVI, and LVEF were the most influential predictors of PV reconnection (Figure 3A). A longer ablation interval and larger LAVI were associated with a higher likelihood of durable isolation at repeat ablation, whereas lower LVEF and older age were linked to reconnection. Pharmacologic variables, including anticoagulant use, hypertension, and rate-control therapy, contributed only modestly to model performance (Figure 3B).

**Figure 3.**
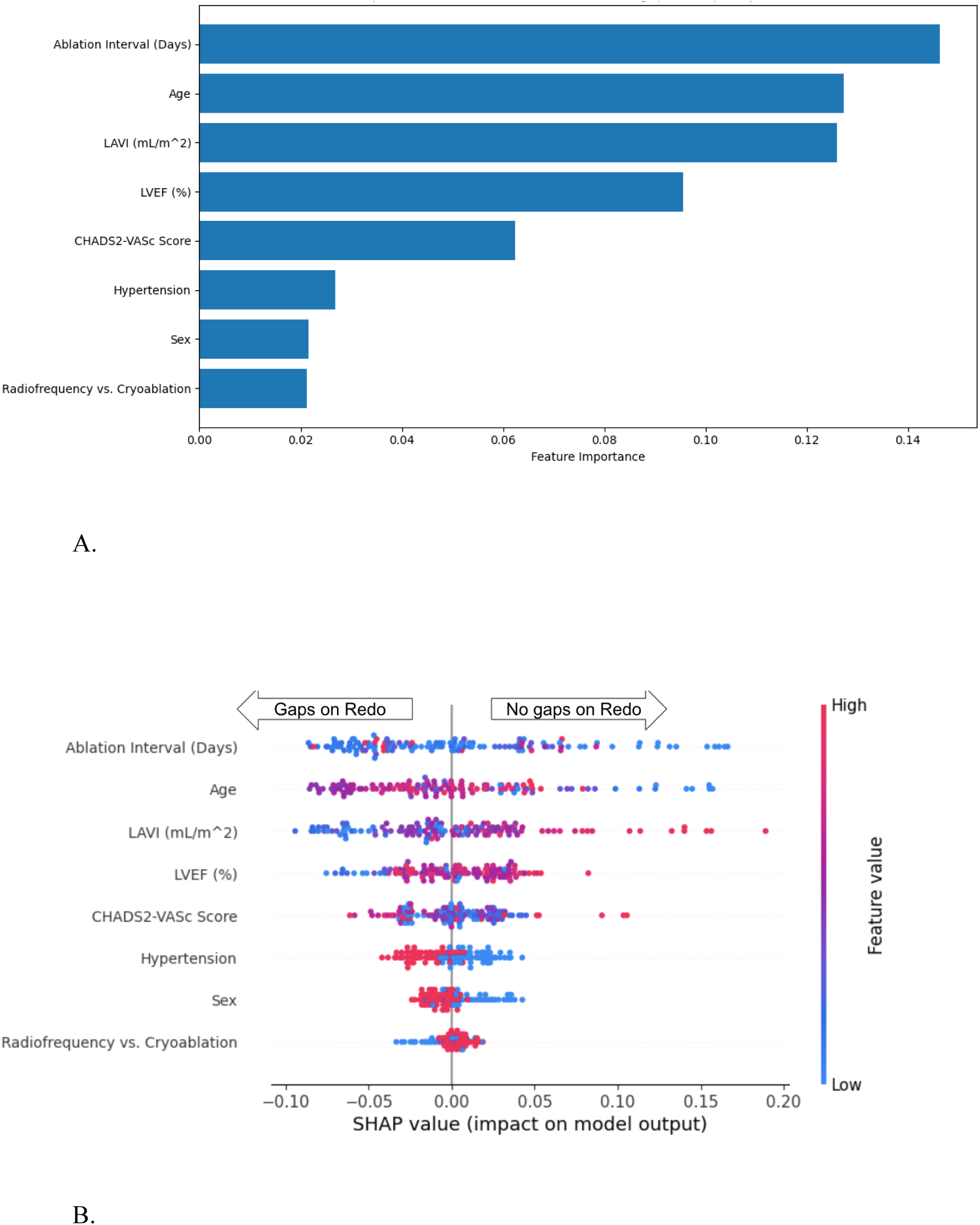
Feature Importance for Predicting Absence of Pulmonary Vein Gaps at Repeat Ablation. Top demographic predictors identified by machine learning models for pulmonary vein (PV) reconnection at repeat procedures. (A) Random Forest feature importance ranking demonstrates that a longer ablation interval, older age, lower left atrial volume index (LAVI), and left ventricular ejection fraction (LVEF) were the strongest contributors. (B) SHAP summary plot showing the direction and magnitude of each feature’s effect on model output. Note: the relationship between feature value and direction may not be linear.

Model interpretability analysis using Gradient-weighted Class Activation Mapping (Grad-CAM) demonstrated that the network consistently focused on the onset and offset of the P-wave as the primary features driving classification (Figure 2B). In contrast, misclassified examples frequently showed attention directed to irrelevant regions, such as the T-wave (Figure 2C). These findings indicate the presence of a detectable biosignature of PVI gaps encoded within P-wave morphology.

In an exploratory analysis, we further assessed whether the initial outpatient ECG obtained prior to the first PVI, when combined with demographic data, could predict failure of the index procedure as defined by the presence of gaps identified during the subsequent redo ablation. This model (DNN-Gap-Initial-Demo) achieved an AUROC of 0.782 (SD 0.036), with a sensitivity of 78.8% (±4.1) and a specificity of 81.2% (±3.8) (Table 1).

## DISCUSSION

This retrospective observational AI study addressed two questions: (i) can a DNN detect a reproducible biosignature of PVI on the surface ECG, and (ii) can pre-procedure ECGs predict residual conduction gaps at redo ablation, and (iii) whether we can predict the failure of the initial PVI procedure. We found that the DNN reliably discriminated pre-versus post-PVI ECGs, with higher performance when inputs were simplified to single-cycle (1–2 s) segments, consistent with reduced input complexity, increased training volume and effective data augmentation. Establishing this capability was a necessary first step, as without a discernible post-PVI biosignature, it would be unlikely for the model to identify more granular endpoints such as gap reconnection, which is the principal clinical objective of this work.

Grad-CAM consistently highlighted the diastolic interval as an important feature for classifying pre-versus post-PVI ECGs, with secondary focus on the P-wave (Figure 2A). One potential explanation is that the model is partially leveraging heart-rate and rhythm-regularity cues encoded in the RR segment - parasympathetic denervation from collateral ablation of ganglionated plexi is known to increase sinus rate and reduce short-term heart-rate variability early after PVI, which would manifest predominantly in diastole.^18,19^ Our data supports this when comparing superimposed pre- and post-ablation ECGs (Supplemental Figure S2). A second possibility is that our windowing strategy (clips centered on the QRS) may bias attention toward inter-beat intervals. These observations suggest that the network is using both morphology (P-wave onset/offset) and timing context (diastolic/RR features) to infer post-ablation status.

The second aim of this study was to determine whether end-to-end deep learning could identify residual conduction gaps from a pre-procedural surface ECG obtained prior to redo ablation. This question is clinically relevant because redo outcomes are generally more favorable when PV reconnection is present, i.e. there is a treatable mechanism, whereas patients with durably isolated PVs, in whom recurrence reflects non-PV triggers or an atrial substrate, tend to have poorer arrhythmia-free survival and limited benefit from further substrate modification.^7–9,20–22^ Prior series have consistently demonstrated that recovered PV conduction is common at redo, and that outcomes are superior when these reconnections can be re-isolated.^5^

In our cohort, a deep learning model trained on pre-procedure ECG segments predicted the presence of PV gaps with an AUROC of 0.819 (SD 0.024), sensitivity of 75.3% (±3.1), and specificity of 78.2% (±2.9), supporting the existence of a biologically plausible ECG biosignature of PV reconnection. Incorporating baseline demographic and clinical features, such as LAVI, interval between procedures, and age, provided a modest but consistent incremental gain (AUROC 0.830 [SD 0.017]), with the best-performing model achieving a sensitivity of 84% and specificity of 72%. These findings underscore that while demographic and structural factors contribute, the surface ECG itself carries the predominant predictive signal for PV gap detection.

Grad-CAM consistently highlighted the onset and offset of the P-wave as the dominant decision features (Figure 2B). This localization is physiologically coherent: P-wave onset predominantly reflects right-atrial activation and conduction via Bachmann’s bundle into the left atrium, whereas the terminal P-wave reflects left-atrial activation. PVI alters left-atrial conduction and PV-related activation fronts, which can shorten, fragment, or otherwise reshape the terminal P-wave component. Prior clinical and computational studies report concordant phenomena, specifically post-ablation changes in P-wave duration and morphology and associations between prolonged filtered P-wave duration/dispersion and AF recurrence, supporting the notion that atrial substrate modification after PVI is encoded in P-wave features on the 12-lead ECG.^23^

Demographic data alone had minimal predictive value for gap recurrence, with models trained solely on clinical variables performing no better than chance. However, when demographic features were combined with surface ECG inputs, model performance improved significantly, which is consistent with the established strength of multimodal models.^24^ Among demographic predictors, the time interval between ablations emerged as the most informative - a longer interval was associated with durable PV isolation, whereas shorter intervals were more likely to reveal reconnection, suggesting that progressive atrial myopathy and non-PV triggers drive later recurrences (Figure 3). Left atrial volume index (LAVI) also emerged as a key marker-larger atria were paradoxically associated with a lower likelihood of PV gaps, consistent with prior studies,^25^ supporting the role of atrial structural disease in extra-PV mechanisms. In contrast, medication use, sex, and ablation modality (radiofrequency vs. cryoablation) did not meaningfully contribute to prediction.

While discrimination was moderate, standalone performance may be insufficient for binary decision-making in all patients. As such, models like ours may have value in risk-stratification of patients to inform treatment selection and patient counseling. Generalizability is limited by the single-center, retrospective design, modest sample size, and potential confounding from heart rate and signal quality. External, prospective validation with vendor-agnostic ECGs is needed, along with cost-effectiveness analyses tied to actionable thresholds (e.g., “no-gap” pathways that avoid invasive remapping). Finally, there is likely an information ceiling in the surface P-wave alone. Multimodal fusion, such as combining ECG with left-atrial fibrosis metrics from late-gadolinium-enhanced cardiac MRI, has shown a strong association with post-ablation outcomes and could meaningfully boost predictive power (e.g., DECAAF and subsequent LGE-MRI studies).^26,27^

Our results extend this literature by demonstrating that end-to-end deep learning can recover this electrophysiologic signal directly from raw 12-lead ECGs and generalize beyond the intraprocedural setting to outpatient recordings, with the strongest performance when comparing ECGs before the first and after the second ablation (AUROC 0.879). Earlier prediction efforts relying on imaging-based or signal-averaged features achieved modest discrimination (e.g., AUC ≈0.6 with 3D LA reconstructions), whereas learning directly from ECG waveforms, potentially integrating subtle timing and morphology cues across leads, appears to yield higher performance.^12^ In parallel, our gap-prediction models using pre-redo ECGs (and incrementally, demographics) achieved AUROC ∼0.82–0.83, aligning with mechanistic expectations that PV reconnection is the dominant cause of recurrence and should imprint on left-atrial activation patterns visible in the P-wave.^28^

## Data Availability

Data can be made available upon direct request.

## NON-STANDARD ABBREVIATIONS AND ACRONYMS

AF: atrial fibrillation
AUC: area under the curve
AUROC: area under the receiver operating characteristic curve
CNN: convolutional neural network
DECAAF: Delayed-Enhancement MRI Determinant of Successful Radiofrequency Catheter Ablation of Atrial Fibrillation
DNN: deep neural network
EARLY-AF: Early Aggressive Invasive Intervention for Atrial Fibrillation
GradCAM: gradient-weighted class activation mapping
IP: intraprocedural
LAVI: left atrial volume index
LGE-MRI: Late gadolinium enhancement magnetic resonance imaging
MLP: multilayer perceptron
OP: outpatient
PV: pulmonary vein
PVI: pulmonary vein isolation
TAILORED-AF: Tailored vs. Anatomical Ablation Strategy for Persistent Atrial Fibrillation

## REFERENCES

1. Andrade JG, Aguilar M, Atzema C, et al. The 2020 Canadian Cardiovascular Society/Canadian Heart Rhythm Society Comprehensive Guidelines for the Management of Atrial Fibrillation. Can J Cardiol. 2020;36(12):1847–1948. doi:10.1016/j.cjca.2020.09.001

2. Andrade JG, Wells GA, Deyell MW, et al. Cryoablation or Drug Therapy for Initial Treatment of Atrial Fibrillation. N Engl J Med. 2021;384(4):305–315. doi:10.1056/NEJMoa2029980

3. Erhard N, Metzner A, Fink T. Late arrhythmia recurrence after atrial fibrillation ablation: incidence, mechanisms and clinical implications. Herzschrittmachertherapie Elektrophysiologie. 2022;33(1):71–76. doi:10.1007/s00399-021-00836-6

4. Ouyang F, Antz M, Ernst S, et al. Recovered Pulmonary Vein Conduction as a Dominant Factor for Recurrent Atrial Tachyarrhythmias After Complete Circular Isolation of the Pulmonary Veins: Lessons From Double Lasso Technique. Circulation. 2005;111(2):127–135. doi:10.1161/01.CIR.0000151289.73085.36

5. Ouyang F, Tilz R, Chun J, et al. Long-Term Results of Catheter Ablation in Paroxysmal Atrial Fibrillation: Lessons From a 5-Year Follow-Up. Circulation. 2010;122(23):2368–2377. doi:10.1161/CIRCULATIONAHA.110.946806

6. Park JW, Yu HT, Kim TH, et al. Mechanisms of Long-Term Recurrence 3 Years After Catheter Ablation of Atrial Fibrillation. JACC Clin Electrophysiol. 2020;6(8):999–1007. doi:10.1016/j.jacep.2020.04.035

7. Deisenhofer I, Albenque JP, Busch S, et al. LB-469805-01 TAILORED CARDIAC ABLATION PROCEDURE FOR PERSISTENT ATRIAL FIBRILLATION GUIDED BY ARTIFICIAL INTELLIGENCE: THE TAILORED-AF RANDOMIZED CLINICAL TRIAL. Heart Rhythm. 2024;21(7):1199. doi:10.1016/j.hrthm.2024.04.025

8. Verma A, Jiang C yang, Betts TR, et al. Approaches to Catheter Ablation for Persistent Atrial Fibrillation. N Engl J Med. 2015;372(19):1812–1822. doi:10.1056/NEJMoa1408288

9. Kistler PM, Chieng D, Sugumar H, et al. Effect of Catheter Ablation Using Pulmonary Vein Isolation With vs Without Posterior Left Atrial Wall Isolation on Atrial Arrhythmia Recurrence in Patients With Persistent Atrial Fibrillation: The CAPLA Randomized Clinical Trial. JAMA. 2023;329(2):127. doi:10.1001/jama.2022.23722

10. Inoue K, Hikoso S, Masuda M, et al. Pulmonary vein isolation alone vs. more extensive ablation with defragmentation and linear ablation of persistent atrial fibrillation: the EARNEST-PVI trial. EP Eur. 2021;23(4):565–574. doi:10.1093/europace/euaa293

11. Alzubaidi L, Zhang J, Humaidi AJ, et al. Review of deep learning: concepts, CNN architectures, challenges, applications, future directions. J Big Data. 2021;8(1). doi:10.1186/s40537-021-00444-8

12. Kim JY, Kim Y, Oh GH, et al. A deep learning model to predict recurrence of atrial fibrillation after pulmonary vein isolation. Int J Arrhythmia. 2020;21(1). doi:10.1186/s42444-020-00027-3

13. Caldwell J, Koppikar S, Barake W, et al. Prolonged P-wave duration is associated with atrial fibrillation recurrence after successful pulmonary vein isolation for paroxysmal atrial fibrillation. J Interv Card Electrophysiol. 2014;39(2):131–138. doi:10.1007/s10840-013-9851-1

14. Saha M, Conte G, Caputo ML, Jacquemet V. Changes in P-wave morphology after pulmonary vein isolation: insights from computer simulations. Europace. Published online 2016. doi:10.1093/europace/euw348

15. Okumura Y, Watanabe I, Ohkubo K, et al. Prediction of the Efficacy of Pulmonary Vein Isolation for the Treatment of Atrial Fibrillation by the Signal-Averaged P-Wave Duration. Pacing Clin Electrophysiol. 2007;30(3):304–313. doi:10.1111/j.1540-8159.2007.00670.x

16. He K, Zhang X, Ren S, Sun J. Deep Residual Learning for Image Recognition. arXiv. Preprint posted online December 10, 2015. doi:10.48550/arXiv.1512.03385

17. Selvaraju RR, Cogswell M, Das A, Vedantam R, Parikh D, Batra D. Grad-CAM: Visual Explanations from Deep Networks via Gradient-based Localization. Int J Comput Vis. 2020;128(2):336–359. doi:10.1007/s11263-019-01228-7

18. Wang X, Hu Z, Yao Y, Maimaitijiang P, Chen A, Zheng L. Changes in heart rate variability parameters following pulsed-field ablation in patients with atrial fibrillation: A systematic review and meta-analysis HRV changes after pulsed-field ablation in AF. IJC Heart Vasc. 2025;60:101766. doi:10.1016/j.ijcha.2025.101766

19. Goff ZD, Laczay B, Yenokyan G, et al. Heart rate increase after pulmonary vein isolation predicts freedom from atrial fibrillation at 1 year. J Cardiovasc Electrophysiol. 2019;30(12):2818–2822. doi:10.1111/jce.14257

20. Darby AE. Recurrent Atrial Fibrillation After Catheter Ablation: Considerations For Repeat Ablation And Strategies To Optimize Success. J Atr Fibrillation. 2016;9(1):1427. doi:10.4022/jafib.1427

21. Schmidt B, Bordignon S, Metzner A, et al. Ablation Strategies for Repeat Procedures in Atrial Fibrillation Recurrences Despite Durable Pulmonary Vein Isolation: The Prospective Randomized ASTRO AF Multicenter Trial. Circulation. 2024;150(25):2007–2018. doi:10.1161/CIRCULATIONAHA.124.069993

22. Valderrábano M, Peterson LE, Swarup V, et al. Effect of Catheter Ablation With Vein of Marshall Ethanol Infusion vs Catheter Ablation Alone on Persistent Atrial Fibrillation: The VENUS Randomized Clinical Trial. JAMA. 2020;324(16):1620. doi:10.1001/jama.2020.16195

23. Masuda M, Inoue K, Iwakura K, et al. Impact of pulmonary vein isolation on atrial late potentials: association with the recurrence of atrial fibrillation. EP Eur. 2013;15(4):501–507. doi:10.1093/europace/eus326

24. Rajpurkar P, Chen E, Banerjee O, Topol EJ. AI in health and medicine. Nat Med. 2022;28(1):31–38. doi:10.1038/s41591-021-01614-0

25. Benali K, Barré V, Hermida A, et al. Recurrences of Atrial Fibrillation Despite Durable Pulmonary Vein Isolation: The PARTY-PVI Study. Circ Arrhythm Electrophysiol. 2023;16(3). doi:10.1161/CIRCEP.122.011354

26. Siebermair J, Kholmovski EG, Marrouche N. Assessment of Left Atrial Fibrosis by Late Gadolinium Enhancement Magnetic Resonance Imaging. JACC Clin Electrophysiol. 2017;3(8):791–802. doi:10.1016/j.jacep.2017.07.004

27. Marrouche NF, Wilber D, Hindricks G, et al. Association of Atrial Tissue Fibrosis Identified by Delayed Enhancement MRI and Atrial Fibrillation Catheter Ablation: The DECAAF Study. JAMA. 2014;311(5):498. doi:10.1001/jama.2014.3

28. Luongo G, Azzolin L, Schuler S, et al. Machine learning enables noninvasive prediction of atrial fibrillation driver location and acute pulmonary vein ablation success using the 12-lead ECG. Cardiovasc Digit Health J. 2021;2(2):126–136. doi:10.1016/j.cvdhj.2021.03.002

